# MethylCog predicts six-year cognitive ability beyond blood-based ADRD biomarkers

**DOI:** 10.64898/2026.05.26.26354133

**Authors:** Deirdre M. O’Shea, Lily Wang, David Lukacsovich, Wei Zhang, James E. Galvin

## Abstract

**INTRODUCTION:** MethylCog is a 29-CpG blood DNA methylation (DNAm) proxy for general cognitive ability (g). Its incremental association with blood biomarkers of Alzheimer’s disease and related dementias (ADRD) and prospective cognitive ability remains unclear.

**METHODS:** In the held-out test set from the original MethylCog study, we tested whether MethylCog explained baseline *g* beyond four ADRD blood biomarkers, and whether it predicted six-year follow-up g beyond baseline *g* and biomarkers.

**RESULTS:** MethylCog showed a stronger age-adjusted association with baseline *g* than individual biomarkers (r=.368 vs absolute r=.083–.162). MethylCog added 10.0% variance beyond all four biomarkers cross-sectionally (p<.001) and predicted six-year follow-up g in the biomarker-adjusted model (β=.108, p=.002). No individual ADRD biomarker independently predicted follow-up g.

**DISCUSSION:** MethylCog may provide cognition-related DNAm information complementary to blood-based ADRD biomarkers.

## 1 Introduction

Blood biomarkers of Alzheimer’s disease and related dementias (ADRD), including phosphorylated tau 181 (pTau181), amyloid-β 42/40 ratio (Aβ42/40), neurofilament light chain (NfL), and glial fibrillary acidic protein (GFAP), index aspects of amyloid/tau pathology, neuroaxonal injury, and astroglial activation [1–4]. However, in population-based samples of older adults, particularly those without dementia, these biomarkers tend to account for a modest proportion of variance in cognitive performance [5, 6]. Blood DNA methylation (DNAm) is influenced by biological aging and cumulative environmental exposures across the life course and has been associated with cognitive outcomes[7–11]. DNAm-derived cognitive indices may therefore capture cognition-related biological variation that is not fully represented by disease-focused blood biomarkers, but relatively few DNAm scores have been developed specifically to index cognition.

In our original study [12], we developed and cross-sectionally validated MethylCog, a sparse 29-CpG elastic net-derived DNAm proxy for general cognitive ability (g), using the Health and Retirement Study Harmonized Cognitive Assessment Protocol (HRS-HCAP)[13]. MethylCog was trained on a comprehensive neuropsychological phenotype rather than a brief telephone-based cognitive measure, showed criterion validity in both a held-out HRS-HCAP test set (N=605) and an independent external cohort (n=112), showed modest associations with mild cognitive impairment (MCI), and captured cognition-related variance distinct from general biological aging, indexed by GrimAge [14]. Exploratory analyses in the external cohort found no significant associations between MethylCog and ADRD blood biomarkers, although the modest sample size and absence of dementia cases limited inference.

Related recent work in the broader HRS Venous Blood Study evaluated a Generation Scotland-derived epigenetic *g* score, computed using genome-wide CpG weights from a BayesR+ model of general cognitive ability [15, 16]. That score was associated with cognitive level measured using the modified Telephone Interview for Cognitive Status (TICS), but was not significantly associated with six-year cognitive decline or incident dementia after covariate adjustment, including adjustment for apolipoprotein E (APOE) ε4 status and blood-based neurodegeneration biomarkers. The present study differs by evaluating MethylCog, a sparse HRS-HCAP-derived DNAm proxy trained on comprehensive neuropsychological general cognitive ability, in the original held-out HRS-HCAP test set using HCAP-derived *g* at baseline and six-year follow-up. At the time of the original MethylCog study, neither the 2022 HRS-HCAP follow-up cognitive data nor the 2016 HRS blood ADRD biomarker data were available for the held-out HRS-HCAP test set. Therefore, the original study could not determine whether MethylCog predicted later HCAP-derived general cognitive ability or whether its cognitive signal overlapped with blood-based ADRD biomarker burden. These data have since become available, allowing the present extension study to test whether MethylCog: (1) accounts for variance in concurrent HCAP-derived *g* beyond ADRD blood biomarkers; and (2) predicts HCAP-derived *g* six years later beyond baseline cognitive level and ADRD blood biomarkers.

## 2. Methods

### 2.1 Participants

Full details of the HRS-HCAP design, neuropsychological battery, DNAm preprocessing, and MethylCog scoring have been described previously in the original MethylCog study [12]. Briefly, the Health and Retirement Study is a nationally representative longitudinal cohort of U.S. adults aged 51 years and older[17]. The original MethylCog study used the 2016 HRS-HCAP wave, a subsample of adults aged 65 years and older, together with whole-blood DNAm data generated from the Illumina Infinium Methylation EPIC array. The present analyses were restricted to the held-out test set from the original study (N = 605). MethylCog scores were computed using the fixed 29-CpG weights and training-set standardization parameters from the original model without modification. The current extension incorporated newly available 2022 HRS-HCAP follow-up cognitive data and newly released 2016 Venous Blood Study neuropathological and supplemental biomarker data, allowing evaluation of whether MethylCog provides cognition-related information beyond blood-based ADRD biomarkers both cross-sectionally and prospectively. Complete data for all four ADRD blood biomarkers were available for 595 of 605 participants. Complete-case analytic samples were smaller after requiring covariate data, with N = 551 for the primary cross-sectional biomarker models and N = 331 for the primary prospective biomarker-adjusted models.

### 2.2 ADRD blood biomarkers

ADRD blood biomarker data were obtained through restricted access from the Health and Retirement Study Venous Blood Study (VBS) 2016 Neuropathological and Supplemental Biomarkers data release, released December 22, 2025 [18]. Blood samples used for biomarker assays were collected as part of the 2016 HRS Venous Blood Study [19], corresponding to the baseline HRS-HCAP wave and the same venous blood collection wave used for DNA methylation profiling. Aβ40, Aβ42, GFAP, and NfL were measured in EDTA plasma using a Quanterix Simoa HD-X analyzer with the Neurology 4-plex assay E. The Aβ42/40 ratio was calculated from measured Aβ42 and Aβ40 values. pTau181 was measured in serum using the Quanterix Simoa pTau-181 V2 assay. All assays were conducted at the University of Minnesota Advanced Research and Diagnostic Laboratory. NfL, GFAP, and pTau181 were natural log-transformed prior to analysis; Aβ42/40 was rank-based inverse-normal transformed for regression analyses.

### 2.3 General cognitive ability

General cognitive ability (*g*) was derived from the HRS-HCAP neuropsychological battery administered at the 2016 baseline and 2022 follow-up waves[13]. Baseline *g* was derived in the original MethylCog study by extracting the first unrotated principal component from z-standardized item-level cognitive test scores, with median imputation for limited item-level missingness[12]. To maintain longitudinal comparability, the 2022 follow-up score was generated by applying the 2016 PCA loadings to standardized 2022 item scores and rescaling the composite to the 2016 metric. Baseline *g* was used as the cross-sectional outcome and as a covariate in prospective models; six-year follow-up g served as the prospective cognitive outcome.

### 2.4 Statistical analyses

Age-adjusted partial correlations characterized associations among baseline *g*, MethylCog, and each blood ADRD biomarker. Cross-sectional analyses used ordinary least squares regression to model baseline *g* among participants with available blood ADRD biomarker data in two steps: (1) age, sex, apolipoprotein E (APOE) ε4 carrier status (any ε4 allele vs none), and all four blood ADRD biomarkers; and (2) the same model with MethylCog added. The increment in adjusted R-squared (ΔAdjR²) attributable to MethylCog was quantified using F-tests for nested model comparisons. Education-adjusted sensitivity analyses added years of education to the corresponding covariate set.

Prospective analyses modeled six-year follow-up *g* using linear regression models that adjusted for baseline *g*. Models followed an equivalent nested structure: (1) baseline *g*, age, sex, and APOE ε4 carrier status; (2) the same model with all four blood ADRD biomarkers added; and (3) each of these models with MethylCog additionally included. This structure allowed direct evaluation of MethylCog’s prospective contribution at each stage of covariate adjustment and tested whether blood ADRD biomarkers added predictive value beyond baseline cognitive level. Education-adjusted sensitivity analyses were conducted in parallel.

To evaluate whether prospective associations were driven by participants with baseline cognitive impairment, sensitivity analyses repeated the prospective models after excluding participants with MCI at baseline. To characterize potential attrition bias, logistic regression was used among participants with complete baseline biomarker and covariate data, including both follow-up completers and non-completers, with follow-up completion status as the binary outcome. Predictors included baseline *g*, MethylCog, age, sex, APOE ε4 carrier status, and all four blood ADRD biomarkers.

All analyses used listwise deletion; N per model reflects the available complete-case sample for that model’s variable set. Analyses were conducted in R version 4.3.

## 3. Results

### 3.1 Sample characteristics

Sample characteristics are presented in Table 1. The baseline test set included 605 participants. Mean age was 75.2 years (SD=7.1), 57.0% were female, and 23.3% met criteria for MCI. Complete data for all four ADRD blood biomarkers were available for 595 participants. Complete-case analytic samples were N=551 for the primary cross-sectional biomarker models and N=331 for the primary prospective biomarker-adjusted models. Six-year follow-up cognitive data were available for 367 participants. Compared with participants without follow-up cognitive data, follow-up completers were younger, more educated, less likely to have MCI at baseline, and had higher baseline *g* and MethylCog scores. They also had lower NfL, GFAP, and pTau181 levels, whereas sex, APOE ε4 carrier status, and Aβ42/40 ratio did not differ by follow-up status.

**Table 1.** Sample characteristics by follow-up status.

| Characteristic | Full test set<br>(N=605) | Follow-up<br>completers<br>(N=367) | Non-completers<br>(N=238) | p value |
| --- | --- | --- | --- | --- |
| Age, mean (SD), years | 75.2 (7.1) | 73.3 (6.1) | 78.1 (7.5) | <.001 |
| Female, n (%) | 345 (57.0) | 209 (56.9) | 136 (57.1) | 1.000 |
| APOE ε4 carrier, n (%) | 131 (23.4)<br>[n=561] | 81 (23.9) [n=339] | 50 (22.5) [n=222] | .785 |
| Education, mean (SD),<br>years | 12.9 (3.0) | 13.2 (3.0) | 12.4 (3.0) | .003 |
| MCI at baseline, n (%) | 141 (23.3) | 60 (16.3) | 81 (34.0) | <.001 |
| MethylCog, mean (SD) | 0.10 (0.97) | 0.21 (0.98) | -0.07 (0.93) | <.001 |
| Baseline *g*, mean (SD) | 0.00 (1.00) | 0.29 (0.85) | -0.43 (1.06) | <.001 |
| Follow-up *g*, mean (SD) | 0.04 (0.97)<br>[n=367] | 0.04 (0.97) | — | — |
| NfL, median (IQR),<br>pg/mL | 22.8 (17.0–33.6)<br>[n=600] | 20.3 (15.2–27.7)<br>[n=362] | 27.7 (20.2–41.7) | <.001 |
| GFAP, median (IQR),<br>pg/mL | 106.1 (73.0–<br>147.5) [n=600] | 94.3 (68.1–131.9)<br>[n=362] | 122.2 (91.6–179.0) | <.001 |
| pTau181, median (IQR),<br>pg/mL | 1.85 (1.32–2.84)<br>[n=596] | 1.67 (1.26–2.52)<br>[n=359] | 2.23 (1.55–3.31)<br>[n=237] | <.001 |
| Aβ42/40 ratio, mean (SD) | 0.066 (0.030)<br>[n=599] | 0.066 (0.015)<br>[n=362] | 0.066 (0.044)<br>[n=237] | .844 |

**Note.** Values are presented as mean (SD), median (IQR), or n (%). Biomarker values are presented on the raw scale. Bracketed n values indicate the number of non-missing observations where this differs from the column total. P values compare follow-up completers and non-completers using t-tests for approximately normally distributed continuous variables, Wilcoxon rank-sum tests for skewed biomarkers, and chi-square or Fisher's exact tests for categorical variables. No p value is reported for follow-up *g* because it was unavailable by definition for non-completers.
**Abbreviations:** APOE, apolipoprotein E; Aβ , amyloid beta; GFAP, glial fibrillary acidic protein; IQR, interquartile range; MCI, mild cognitive impairment; NfL, neurofilament light chain; pTau181, phosphorylated tau at threonine 181; SD, standard deviation.

### 3.2 Attrition analysis

In a multivariable logistic regression including both follow-up completers and non-completers with complete baseline biomarker and covariate data, baseline *g* (OR=1.96, p<.001) and age (OR=0.95, p=.005) were the only significant independent predictors of follow-up completion. MethylCog did not independently predict completion (OR=0.97, p=.809), nor did any individual blood biomarker.

### 3.3 Bivariate associations among MethylCog, *g*, and ADRD blood biomarkers

Age-adjusted partial correlations are shown in **Table 2**. MethylCog showed the strongest association with baseline *g* (r=.368, p<.001) among the molecular measures examined. Age-adjusted correlations between individual ADRD blood biomarkers and baseline *g* were smaller in magnitude: GFAP (r=–.162, p<.001), NfL (r=–.133, p=.001), pTau181 (r=–.111, p=.007), and Aβ42/40 (r=–.083, p=.042). MethylCog’s age-adjusted correlations with the biomarkers were small: NfL (r=–.080, p=.049), Aβ42/40 (r=–.110, p=.007), GFAP (r=–.072, p=.080), and pTau181 (r=–.012, p=.772). Among the biomarkers, NfL and GFAP were substantially intercorrelated (r=.487, p<.001), while Aβ42/40 was largely independent of the neurodegeneration markers.

**Table 2.**
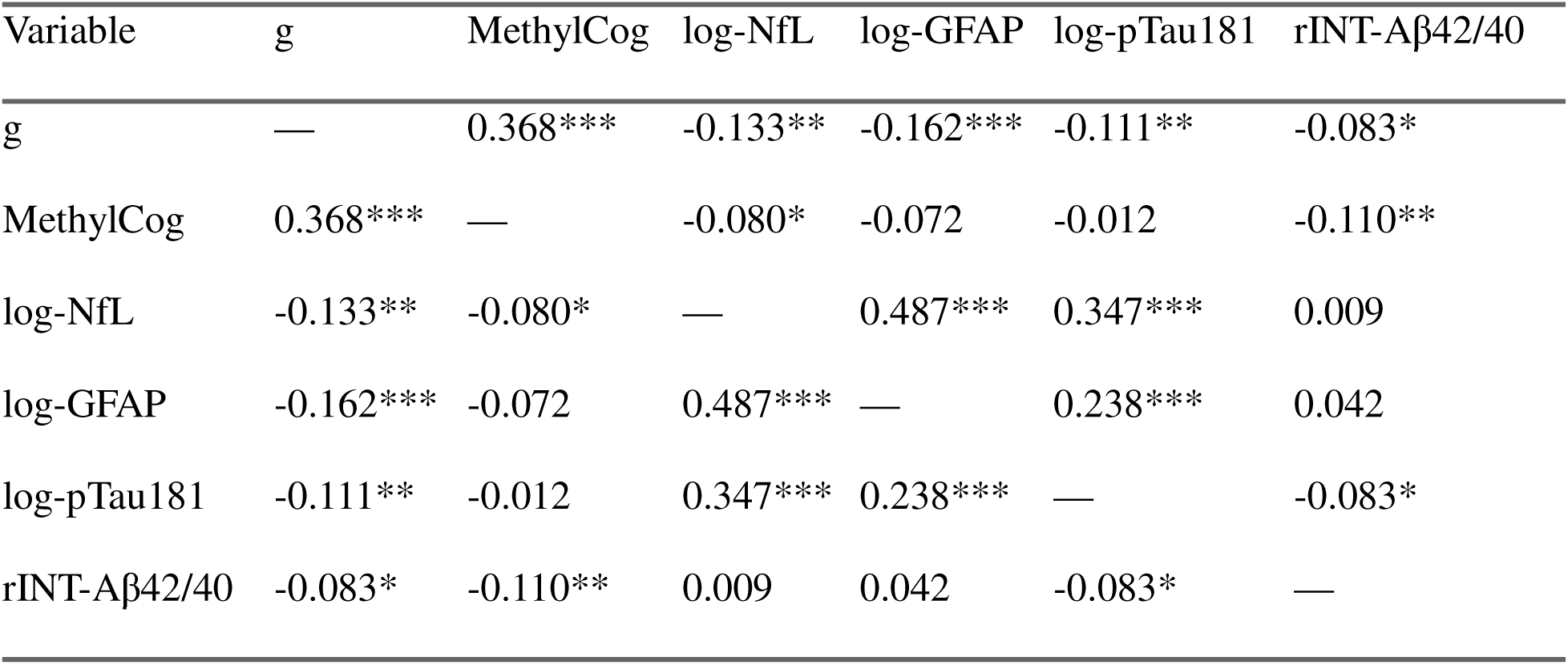
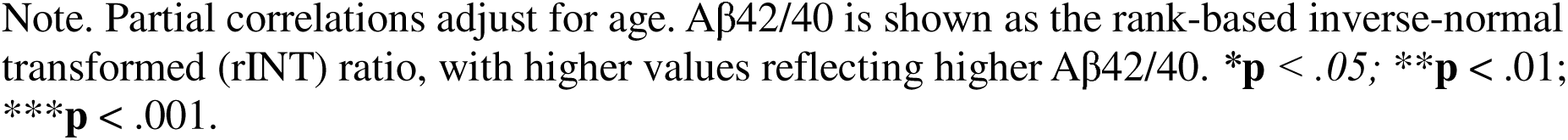
Age-adjusted partial correlations among general cognitive ability (*g*), MethylCog, and blood-based ADRD biomarkers.

| Variable | *g* | MethylCog | log-NfL | log-GFAP | log-pTau181 | rINT-Aβ42/40 |
| --- | --- | --- | --- | --- | --- | --- |
| *g* | — | 0.368*** | -0.133** | -0.162*** | -0.111** | -0.083* |
| MethylCog | 0.368*** | — | -0.080* | -0.072 | -0.012 | -0.110** |
| log-NfL | -0.133** | -0.080* | — | 0.487*** | 0.347*** | 0.009 |
| log-GFAP | -0.162*** | -0.072 | 0.487*** | — | 0.238*** | 0.042 |
| log-pTau181 | -0.111** | -0.012 | 0.347*** | 0.238*** | — | -0.083* |
| rINT-Aβ42/40 | -0.083* | -0.110** | 0.009 | 0.042 | -0.083* | — |

Note. Partial correlations adjust for age. Aβ 42/40 is shown as the rank-based inverse-normal transformed (rINT) ratio, with higher values reflecting higher Aβ 42/40. \* $p < .05$ ; \*\* $p < .01$ ; \*\*\* $p < .001$ .

### 3.4 Cross-sectional: MethylCog beyond blood ADRD biomarkers

Cross-sectional models tested whether MethylCog explained variance in *g* beyond demographic covariates, APOE ε4 status, and the ADRD blood biomarker panel. Results are shown in Table 3. In the model adjusted for age, sex, APOE ε4, and all four blood biomarkers, individual biomarker effects were small after simultaneous entry. Adding MethylCog increased AdjR² from .211 to .311 (ΔAdjR² = .100, F change = 79.73, p < .001; β = 0.339, 95% CI: 0.264–0.413). The increment remained significant in the education-adjusted sensitivity analysis, though attenuated (ΔAdjR² = .023, p < .001; β = 0.177, 95% CI: 0.108–0.247), with GFAP and pTau181 emerging as significant predictors only when education was included in the model.

**Table 3.**
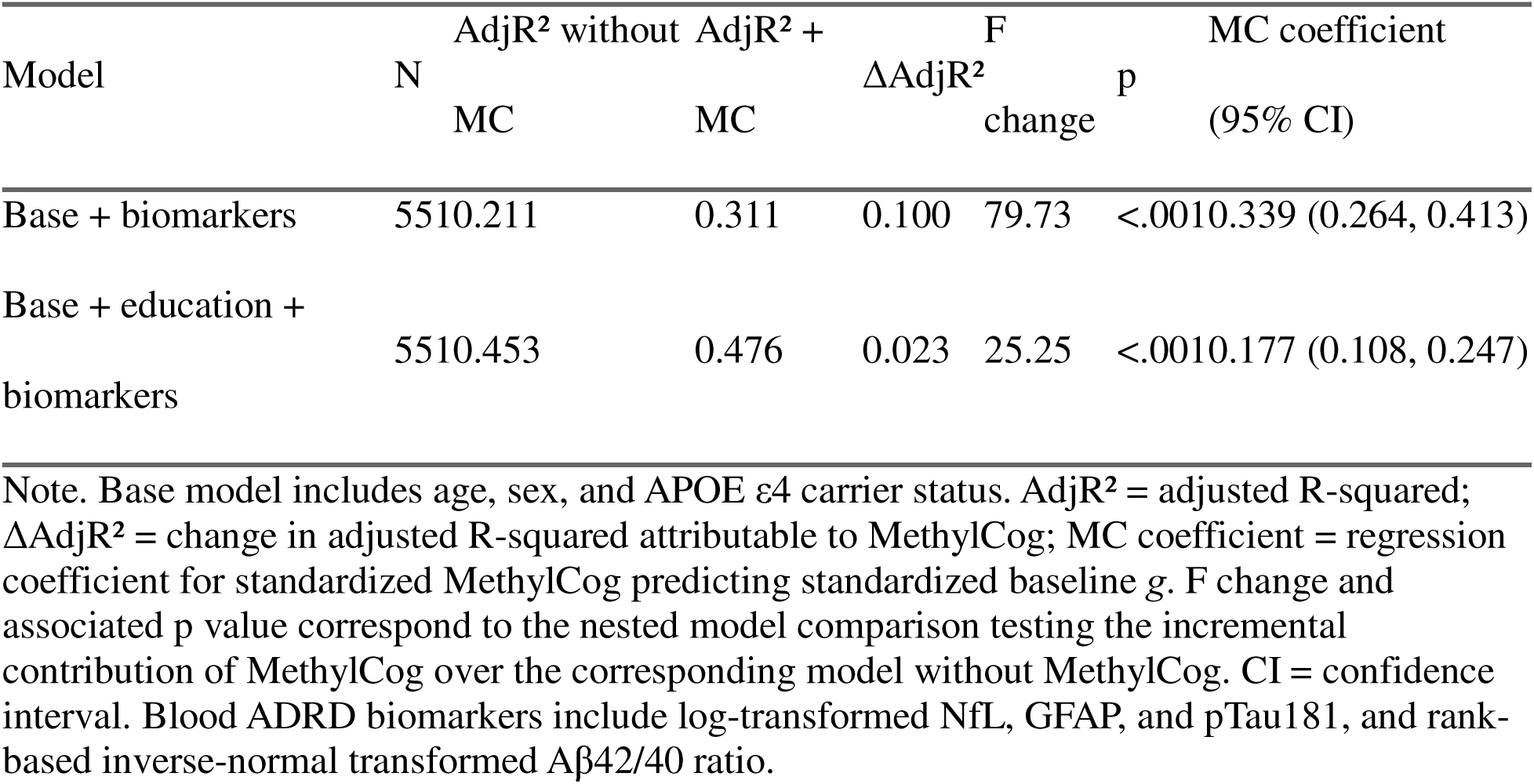
Cross-sectional nested models: incremental effect of MethylCog (MC) beyond blood ADRD biomarkers in predicting baseline general cognitive ability (g).

### 3.5 Prospective: MethylCog predicts g at six-year follow-up

Prospective model results are shown in Table 4. After adjusting for baseline *g*, age, sex, and APOE ε4 carrier status, each 1-SD higher MethylCog score was associated with 0.101 SD higher *g* at six-year follow-up (β = 0.101, 95% CI: 0.034–0.167, p = .003; N = 339, ΔAdjR² = .008). This association remained significant with education additionally included (β = 0.094, p = .007). When all four blood biomarkers were added to the model alongside baseline *g* and demographic covariates, none of the individual ADRD blood biomarkers were significantly associated with follow-up *g* (all p > .14). MethylCog remained a statistically significant predictor in the primary biomarker-adjusted model, such that each 1-SD higher MethylCog score was associated with 0.108 SD higher *g* at follow-up (β = 0.108, 95% CI: 0.040–0.175, p = .002; N = 331, ΔAdjR² = .009). Education-adjusted sensitivity analyses yielded comparable findings (β = 0.098, p = .005). Sensitivity analyses restricted to participants who were cognitively normal at baseline also yielded a similar pattern, with MethylCog remaining associated with six-year follow-up *g* in the model adjusted for baseline *g*, demographics, education, and all four ADRD blood biomarkers (β = 0.110, 95% CI: 0.040–0.179, p = .002; N = 280). The adjusted association between MethylCog and six-year follow-up *g* is shown in **Figure 1**.

**Figure 1.** Association between baseline MethylCog and general cognitive ability (*g*) at six-year follow-up. Both variables were residualized for baseline *g*, age, sex, APOE ε4 carrier status, log-NfL, log-GFAP, log-pTau181, and rINT-Aβ42/40 in the primary biomarker-adjusted analytic sample (N = 331). The solid line represents the ordinary least squares fit, and shading indicates the 95% confidence band. The corresponding prospective model estimate was β = 0.108, 95% CI: 0.040–0.175, p = .002.

**Table 4.** Prospective nested models: MethylCog predicting general cognitive ability at six-year follow-up.

| Model | Specification | N | AdjR<sup>2</sup> without MC | AdjR<sup>2</sup> + MC | ΔAdjR<sup>2</sup> | F change | p | MC coefficient (95% CI) |
| --- | --- | --- | --- | --- | --- | --- | --- | --- |
| Base | Without biomarkers | 339 | 0.664 | 0.672 | 0.008 | 8.75 | 0.003 | 0.101 (0.034, 0.167) |
| Base | With biomarkers | 331 | 0.667 | 0.676 | 0.009 | 9.86 | 0.002 | 0.108 (0.040, 0.175) |
| Base + | Without | 339 | 0.665 | 0.672 | 0.006 | 7.30 | 0.007 | 0.094 (0.026, |

| Model | Specification | N | AdjR<sup>2</sup><br>without<br>MC | AdjR<sup>2</sup> +<br>MC | ΔAdjR<sup>2</sup> | F<br>change | p | MC coefficient<br>(95% CI) |
| --- | --- | --- | --- | --- | --- | --- | --- | --- |
| education | biomarkers |  |  |  |  |  |  | 0.163) |
| Base +<br>education | With<br>biomarkers | 331 | 0.670 | 0.677 | 0.007 | 7.83 | 0.005 | 0.098 (0.029,<br>0.167) |

Note. Base model includes baseline g, age, sex, and APOE ε4 carrier status. AdjR<sup>2</sup> = adjusted R-squared; $\Delta$ AdjR<sup>2</sup> = change in adjusted R-squared attributable to MethylCog; MC coefficient = regression coefficient for standardized MethylCog predicting standardized six-year follow-up g. F change and associated p value correspond to the nested model comparison testing the incremental contribution of MethylCog over the corresponding model without MethylCog. CI = confidence interval.

## 4 Discussion

Using newly released blood biomarker and six-year follow-up cognitive data from the HRS-HCAP test set, this extension study evaluated whether MethylCog provides cognition-related information beyond blood-based ADRD biomarkers. MethylCog accounted for additional variance in baseline *g* beyond NfL, GFAP, pTau181, and Aβ42/40, and remained associated with six-year follow-up g after adjustment for baseline *g* and demographic covariates. In contrast, individual ADRD blood biomarkers showed modest associations with baseline *g*, and biomarker effects were not retained prospectively after accounting for baseline *g*. These findings suggest that MethylCog indexes cognition-related DNAm variation that is not fully explained by currently available blood-based ADRD biomarkers.

Overall, these findings are consistent with the broader interpretation that cognitive performance in population-based older adult samples reflects more than measurable ADRD biomarker burden. Blood biomarkers provide increasingly informative indices of amyloid/tau-related processes, neuroaxonal injury, and astroglial activation, and have demonstrated prognostic value for dementia risk in community-based cohorts[6, 20]. However, their associations with continuous cognitive performance may be modest or inconsistent in samples that are not enriched for dementia or elevated biomarker burden [6]. In the present sample, individual ADRD biomarker associations with cognition were small, and biomarker effects were not retained prospectively after accounting for baseline *g*. This does not diminish the value of ADRD blood biomarkers for detecting disease-related biological processes; rather, it suggests that these markers explain only part of the variance in cognitive performance in a predominantly normal-to-MCI population-based sample.

The current findings extend the original MethylCog study in two important ways. First, they provide prospective evidence that a fixed baseline MethylCog score predicts general cognitive ability six years later, beyond participants’ starting cognitive level. This is an important distinction because a DNAm score may correlate with cognition cross-sectionally without carrying predictive information over time. Notably, the prospective association persisted after excluding participants with baseline MCI, suggesting that the finding was not driven solely by individuals who were already clinically impaired at baseline. Second, the present analyses provide a larger test of the exploratory biomarker findings from the original external validation cohort, where MethylCog showed no significant associations with ADRD plasma biomarkers but inference was limited by the modest sample size and absence of dementia cases. In the current HRS-HCAP test-set sample, age-adjusted correlations between MethylCog and individual blood ADRD biomarkers were small, suggesting that MethylCog is not simply a proxy for measured biomarker burden.

These findings also complement recent HRS work evaluating a Generation Scotland-derived epigenetic *g* score. That study found that epigenetic *g* was associated with modified TICS-based cognitive level but showed limited evidence for prediction of six-year cognitive decline or incident dementia after covariate adjustment [15, 16]. The present study differs by evaluating MethylCog, a sparse 29-CpG score trained on comprehensive HRS-HCAP general cognitive ability, and by testing prediction of follow-up HCAP-derived *g* within the original held-out test set. Direct comparison of MethylCog with epigenetic *g* will be important in future work.

Conceptually, these findings align with reserve and resilience frameworks in which cognitive performance reflects both disease burden and individual differences in cognitive capacity, brain maintenance, and life-course influences [21–24]. MethylCog should not be interpreted as an ADRD diagnostic biomarker. It was developed to index general cognitive ability, not amyloid, tau, neurodegeneration, or diagnostic status. Its potential value may therefore lie in capturing cognition-related biological variation that complements pathology-focused biomarkers. This distinction may be especially relevant in population-based studies, retrospective biospecimen analyses, cohorts where direct cognitive assessment is incomplete or unavailable, and clinical trial recruitment contexts where MethylCog could eventually complement cognitive testing and ADRD biomarkers for participant stratification or enrichment.

The attenuation of MethylCog’s cross-sectional association after education adjustment is also informative. Educational attainment is strongly related to cognitive performance and reflects, in part, cumulative socioeconomic, developmental, and environmental exposures [25, 26], factors shown to influence DNAm [10, 27, 28]. Therefore, some overlap between MethylCog and education-related variance is expected. Importantly, MethylCog remained associated with cognition after adjustment for education and ADRD blood biomarkers, suggesting that it captures cognition-related epigenetic information not fully accounted for by educational attainment alone.

Several limitations should be noted. These analyses use the same held-out HRS-HCAP test set from the original MethylCog study and therefore constitute an internal prospective extension rather than independent external replication. The 39% absence of follow-up cognitive data introduced potential attrition and survivor-selection bias: participants with follow-up data were younger and had higher baseline cognition. However, MethylCog was not associated with follow-up completion, and the strongest predictors of completion, baseline *g* and age, were included as covariates in all prospective models. Residual attrition bias cannot be ruled out. The prospective sample was also modest in size, limiting generalizability and power for interaction analyses. Finally, MethylCog was measured at a single time point. Repeated DNAm assessments are needed to determine whether MethylCog primarily reflects stable individual differences in cognitive capacity or dynamic methylation changes associated with cognitive trajectories.

Taken together, this extension study suggests that MethylCog indexes cognition-related DNAm variation that is not fully explained by currently available blood-based ADRD biomarkers and provides independent prospective information over a six-year interval. Future studies should include independent replication in cohorts with longitudinal cognitive and biomarker data, evaluation of MethylCog in samples with greater pathological burden where biomarker–cognition associations may be stronger, and testing whether MethylCog stratifies risk for incident MCI and dementia in population-based prospective designs.

## Acknowledgments

The authors thank the participants and staff of the Health and Retirement Study-Harmonized Cognitive Assessment Protocol (HRS-HCAP) for their contributions to this research.

## Ethics Statement

The Health and Retirement Study is approved by the Institutional Review Board of the University of Michigan. The present secondary analysis of Health and Retirement Study and NIAGADS data was approved by the Institutional Review Board of the University of Miami. DNA methylation data were accessed through NIAGADS under an approved Data Use Agreement.

## Consent Statement

All participants provided written informed consent prior to participation in the study under a Certificate of Confidentiality from the National Institutes of Health.

## Data Availability Statement

Data used in this study are available through the Health and Retirement Study Data used in this study are available through the Health and Retirement Study (HRS) and the National Institute on Aging Genetics of Alzheimer’s Disease Data Storage Site (NIAGADS). Public-use HRS demographic and cognitive data are available through the HRS website(https://hrs.isr.umich.edu). HRS-HCAP, APOE genotype, Venous Blood Study biomarker data, and HRS linkage/cross-reference files require restricted or sensitive data access approval through HRS https://hrsdata.isr.umich.edu/data-products/sensitive-health. DNA methylation data are available through NIAGADS under accession NG00153 after an approved data access request and Data Use Agreement (https://dss.niagads.org/datasets/ng00153/). Code and scoring files for MethylCog, including the 29-CpG weights, CpG-level training-set scaling parameters, score-level scaling parameters, synthetic example data, expected output, and example R implementation, are publicly available through GitHub and archived on Zenodo, version v1.0.1 (DOI: 10.5281/zenodo.20430921). Individual-level HRS, HBI, DNA methylation, cognitive, biomarker, neuroimaging, APOE, clinical, and diagnostic data are not publicly shared because they contain restricted participant-level information.

